# Murine Fetal Tracheal Occlusion Increases Lung Basal Cells via Increased Yap Signaling

**DOI:** 10.1101/2021.09.17.21263741

**Authors:** Vincent Serapiglia, Chad Stephens, Rashika Joshi, Emrah Aydin, Marc Oria, Mario Marotta, Jose L. Peiro, Brian M. Varisco

**Author notes:** Corresponding author (Brian Varisco). Authors contributed equally to the development of the manuscript (Vincent Serapiglia, Chad Stephens). Funding: Varisco: R01HL141229.

## Abstract

Fetal endoscopic tracheal occlusion (FETO) is an emerging surgical therapy for congenital diaphragmatic hernia (CDH). Ovine and rabbit data suggested altered lung epithelial cell populations after TO with transcriptomic signatures implicating basal cells. To test this hypothesis, we deconvolved mRNA-seq data and used quantitative image analysis in fetal rabbit lungs to showed increased basal cells and reduced ciliated cells after TO. In a fetal mouse TO model, flow cytometry showed increased basal cells, and immunohistochemistry demonstrated basal cell extension to the subpleura. Nuclear yap, a known regulator of basal cell fate, was increased in TO lung, and Yap ablation on the lung epithelium abrogated TO-mediated basal cell expansion. mRNA-seq of TO lung showed increased activity of downstream Yap genes. Human lung specimens with congenital and fetal endoscopic tracheal occlusion had clusters of subpleural basal cell that were not present in control. TO increases lung epithelial cell nuclear Yap leading to basal cell expansion.

## INTRODUCTION

Congenital diaphragmatic hernia (CDH) results from the failed fusion of the pleuroperitoneal folds in the 6^th^ to 8^th^ week of gestation in the human and is present in 1:2000 births (Al-Maary et al., 2016). In CDH, abdominal contents herniate into the thorax causing ipsilateral and contralateral lung hypoplasia and pulmonary vascular pruning. The morbidity and mortality associated with CDH is principally determined by the size of the defect and the laterality, with right sided hernia experiencing worse outcomes. The most severe defects account for 20-35% of all CDH and have an associated at least 50% mortality with severe respiratory morbidity in survivors (Coughlin et al., 2015). For this reason, a fetal surgical technique has been developed to improve fetal lung growth in CDH.

Fetal endotracheal occlusion (FETO) was first described in animal models of CDH by Harrison *et al*. in 1980 (Harrison et al., 1980), and human clinical trials have been ongoing for 35 years (Al-Maary et al., 2016). Typically, FETO involves placement of an endoluminal tracheal balloon at 26-29 weeks’ gestation with fetoscopic removal at 34 weeks. Tracheal occlusion prevents the egress of epithelial secretions and causes lung expansion and growth. A meta-analysis of multiple small, randomized trials indicated a survival benefit of FETO but suggested continued respiratory morbidity (Al-Maary et al., 2016). Larger randomized trials are ongoing. Studies regarding the effects of CDH and FETO on lung cell function, there are mixed reports of CDH impact on alveolar type II cell function (Boucherat et al., 2007; Chapin et al., 2005; Davey et al., 2005), and a report that CDH reduces and TO increases TGF-β signaling and lung elastin synthesis (Vuckovic et al., 2016). Two whole lung gene expression profiling manuscripts using a rabbit model reported complimentary findings of decreased cell proliferation in CDH with restoration or excessive restoration of cell division with FETO (Engels et al., 2016; Varisco et al., 2016). Our group reported preferential proliferation in the epithelial compartment in FETO that was potentially EGFR-related (Varisco et al., 2016). Proteomic analysis of tracheal fluid in an ovine model of CDH and FETO found PI3K and mTOR signaling was increased with reduced abundance of ciliated cells in FETO (Peiro et al., 2018). Adding to the complexity of animal models is a recent finding that in the rabbit model of CDH and FETO there are significant metabolic differences between and within lung lobes (Dobrinskikh et al., 2020). A barrier to understanding the mechanistic underpinning of changes in CDH and TO has been the lack of a mouse TO model with high fetal survival rates. This obstacle has been recently overcome with the development of a transuterine mouse TO model (Aydin et al., 2018; Aydın et al., 2021a).

The goal of this study was to characterize the impact of fetal tracheal occlusion on the lung epithelium. To do so, we first performed a *post hoc* analysis of cell-specific genes derived from the left lower lobes of fetal rabbits with CDH and/or FETO and then validated these findings by histology of the left upper lobes. After identifying basal cells as the most dysregulated cell type, we analyzed the whole lungs of mice in a mouse model of FETO to identify these same changes and identify Hippo/Yap as a key transcriptional regulator using a lung epithelial cell-specific knockout approach. We lastly show that the lungs of human neonates with congenital tracheal occlusion have clusters of basal cells in the distal lung that are not present in the lung of control infants.

## MATERIALS AND METHODS

### Human Tissue Use

Archived human tissues were used with a waiver from the Cincinnati Children’s Hospital Institutional Review Board (2016-9641) and from the Vall Hebron Institute Research.

### Animal Care and Maintenance

All animal use was approved by the Cincinnati Children’s Hospital IACUC (2019-0034). Timed pregnant C57BL/6 mice were housed in a barrier facility with chow and water *ad lib* and 12-hour day-night cycles. Timed pregnant New Zealand white rabbits were purchased from Charles River (Wilmington, MA, USA) and similarly housed.

### Rabbit CDH and TO Model

The fetal rabbit CDH and TO model has been previously described (Varisco et al., 2016). In brief, after anesthesia of the pregnant rabbit, in fetuses, a left sided diaphragmatic defect is created at E25, TO is performed at E27, and fetuses were collected at E30.

### Mouse TO Model

The transuterine mouse TO model has been previously described (Aydin et al., 2018; Aydın et al., 2021a). Briefly, E16.5 C57BL/6 mice were anesthetized and fetuses exposed by midline laparotomy. Anterior-facing fetuses had ligation of the trachea and one carotid artery and jugular vein by passing a non-cutting needle with 6-0 silk through the uterine wall and around those structures. 2 mg progesterone was administered intramuscularly at E17.5 to prevent spontaneous abortion. At E18.5, pregnant mice were anesthetized, fetuses collected, and both fetuses and dam sacrificed.

### Tissue Processing

Lung tissue was snap frozen on dry ice and stored at -80°C. In rabbits the lower left lobe and in mice the entire lung was homogenized and RNA extracted using RNEasy kits (Qiagen). Rabbit lung histology was obtained from frozen left upper lung tissue in OCT (Tissue-Tek) and cryostat sectioning. Entire fetal mouse lungs were fixed in 4% paraformaldehyde in PBS, dehydrated, paraffinized, and sections obtained. For lung lysates, frozen lung tissue was homogenized in RIPA buffer.

### Immunofluorescence (IF) and Immunohistochemistry (IHC)

For IHC, mouse lung sections were stained using anti-NGFR (Abcam, ab8874, 1:1000), anti-Yap (Cell Signaling, 4912S, 1:1000), anti-phospho-YAP(Cell Signaling, 13008S, 1:1000), and anti-Ki67 (AbCam, ab16667, 1:1000) antibodies with ABC Vectastain kit (Vector Laboratories). For IF, rabbit lung sections were stained with pro-Surfactant Protein C (raised in guinea pig, gift of Jeffrey Whitsett, 1:200), anti-CGRP (Abcam, ab360001, 1:200), anti-acetylated alpha tubulin (Milipore, MABT868, 1:200), anti-Cytokeratin 14 (Abcam, ab7800, 1:200), anti-HopX(Abcam, ab57832, 1:200), anti-Mucin 5AC (Abcam, ab3649, 1:200), anti-Scgb1a1 (raised in guinea pig, gift of Jeffrey Whitsett, 1:200). For IF, human lung specimens were stained with anti-Yap (Cell Signaling, 4912S, 1:200) and anti-Keratin-5 (AbCam, ab53121, 1:200).

### Quantitative Image Analysis

For rabbit lung specimens, images of the entire lung lobe at 10X magnification were generated be tile scanning with auto-focusing of each frame at 10X magnification on a Nikon SpectraX microscope. For each analyte, a negative control image was used to determine an intensity threshold and size thresholds determined based on the marker (nuclear vs. cytoplasmic, vs. apical membrane). Non-lung material was manually excluded, and the percentage of cells expressing each analyte determined as nuclei associated with the marker divided by total nuclei. These percentages were then compared by Wilcoxon rank-sum test with Kruskal-Wallis *post hoc* comparisons.

### Western Blot

Mouse lung homogenates were electrophoretically separated and transferred to PVDF membranes and probed for connective tissue growth factor (AbCam, ab6992, 1:1000), cytokeratin-14 (Abcam, ab7800, 1:1000), and β-actin (Abcam, ab184092, 1:1000).

### Flow Cytometry

E18.5 mouse lungs were collected and single cell suspension created as previously described (Joshi et al., 2016). Cells were stained for NGFR (Abcam, ab8874, 1:200) and EpCam (eBioscience, 47-579180, 1:200) antibodies using Zombie Red (BioLegend, 423109) viability dye for dead cell exclusion on a FACSCanto cell (BD Biosciences) flow cytometry device and using FlowJo v10.7.1 software for quantification.

### PCR

The quantities of lung cell-specific mRNAs in Rabbit lung was quantified by SybrGreen (Qiagen, 204143) RT-PCR using the oligonucleotides in Supplemental Table 3.

### mRNA-Seq and Bioinformatic Analysis

Lung cell-specific mRNAs were extracted from LungGENS E16.5, E18.5, PND1, PND3, PND7, and PND28 profiles (Du et al., 2015) and the FKPM values of these genes extracted from a previously published rabbit CDH and TO dataset (GSE84811). The median FKPM per group was used for comparison of cell-specific mRNAs. For mouse TO experiments, RNA from control and TO mouse lungs was barcoded using NexteraXT and Illumina oligos and sequenced at a depth of 10 million reads per sample with 150 base-pair, paired end sequencing on a Novaseq sequencer (Illumina). Reads were aligned to GRCm38 using STAR (Dobin et al., 2013) and count files imported into DESeq2 (Love et al., 2014, p. 2) using a threshold of 5 reads per gene in all samples and differential expression by group determined. Deferentially expressed genes (DEGs) with an adjusted p-value of <0.1 and fold change of 2 or greater were exported into ToppGene (Chen et al., 2009) and Ingenuity Pathway Analysis (IPA) (Krämer et al., 2014) for gene set enrichment analysis (GSEA).

### Statistical Analysis

R version 4.0.2 (R Core Team, 2019), ggpubr (“ggpubr,” n.d.), and rstatix (Kassambara, 2020) were used for graphical and statistical comparisons using Kruskal-Wallis with Dunn’s *post hoc* test using Bonferroni correction for multiple comparisons with p-values of less than 0.05 considered significant. Line and whisker plots indicate medians and 25th and 75th percentiles.

## RESULTS

### Increased Numbers of Lung Basal Cells in Rabbit FETO Model

To begin identifying the lung cells most impacted by FETO, we generated lists of cell-specific mRNAs (Supplemental Table 1) using the Lung Gene Expression Analysis database (Du et al., 2015). Normalizing the FKPM values to of fetal rabbits undergoing fetal creation of diaphragmatic hernia (CDH), FETO, or both (CDH/FETO) to control, we identified increased relative expression of basal cell-specific mRNAs, and decreased relative expression of club, ciliated, and goblet cell mRNAs in FETO and CDH/FETO lungs. There were no substantial differences in the relative abundances of alveolar type 1 (AT-1), AT-2, lipofibroblast, matrix fibroblast, myofibroblast, epithelial, or myeloid cell mRNAs (Figure 1A). As the focus of this project was TO, we extracted the 100 genes with adjusted p-value <0.1 and 1.5-fold increased or decreased expression in control vs TO and performed GSEA. Cell-cycle-related processes were enriched, and basal cell genes were over-represented (Table 1). Quantification of cell-specific mRNAs was consistent with these bioinformatic findings (Supplemental Figure 1). On the protein level, cell-specific markers identified an increased number of basal cells in TO lung (Figure 1B-D) and reduced numbers of AT1 cells in CDH, TO, and CDH/TO lung compared to control (Figure 1E-G). This analysis of lung tissue from a previously published rabbit CDH/TO study shows that TO lung has increased numbers of basal cells compared to control.

**Figure 1:**
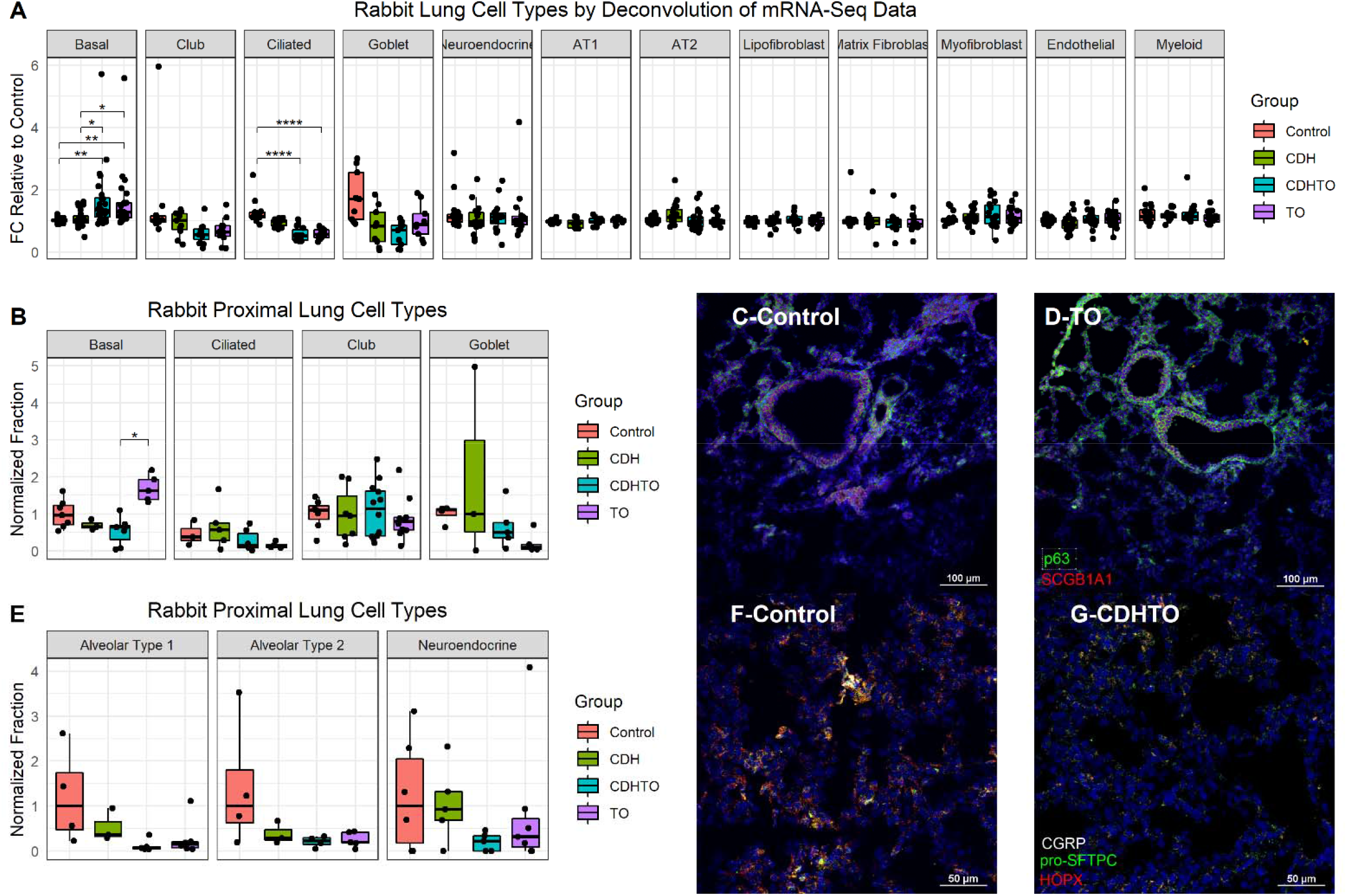
Increased Basal Cells in Rabbit Model of Fetal Tracheal Occlusion. (A) Lung cell-specific mRNAs were identified and the relative expression of each in rabbit fetuses with sham surgery, diaphragmatic hernia creation (CDH), tracheal occlusion (TO) or both (CDHTO). Each data point represents the relative expression of one gene per group. This analysis identified a relative increase in basal cell and reduction in ciliated cell-associated genes. *p<0.05, **p<0.01, ****p<0.0001 by Dunn’s *post hoc* test with Bonferroni correction. (B) Immunostaining and quantitative image analysis upper left lobe rabbit lung showed a relative increase in the number of basal cells as a fraction of total cells in TO lung. (C) The basal cell master regulator p63 was present in conducting airways of control rabbit fetuses largely below SCGB1A1-positive club cells. (D) The relative abundance of p63-positive cells was increased in the lungs of TO rabbit fetuses. (E) Apparent reductions of both alveolar typ1 and alveolar type 2 cells were not statistically significant. (F) Immunostaining for the alveolar type 2 cell marker pro-Surfactant Protein C (pro-SFTPC) and the alveolar type 1 cell marker HOPX in sham-treated fetal rabbit lung. (G) CDHTO lung appeared to have reductions in both markers.

**Table 1.** Gene Set Enrichment Analysis of Fetal Rabbit TO vs. Control.

| <u>GO Molecular Function</u> | <u>GO Biological Process</u> | <u>TopCell Atlas</u> |
| --- | --- | --- |
| DNA replication origin binding | pre-replicative complex assembly | Cycling Basal Cell (Mouse Trachea) |
| calcium ion binding | pre-replicative complex assembly involved in cell cycle DNA replication | Basal Cell (Mouse Trachea) |
| extracellular matrix structural constituent | regulation of cell population proliferation | Dividing T-cell (Human Spleen) |
| glycosaminoglycan binding | double-strand break repair via break-induced replication | Highly activated T cell (Human Ileum) |
| single-stranded DNA binding | circulatory system development | Cycling Basal Cell (Human Nasal Brushing) |
| DNA-dependent ATPase activity | tube development | AT1-AT2 progenitors (Human Lung) |
| drug binding | vasculature development | Spleen Cells (Human) |
| cell adhesion molecule binding | cell adhesion | Matrix Fibroblast (Mouse Lung) |
| DNA helicase activity | biological adhesion | Dividing Monocyte (Human Lung) |
| single-stranded DNA helicase activity | urogenital system development | B-cells (Human Spleen) |

### Increased Number of Basal Cells After Fetal Mouse Tracheal Occlusion

We next used a recently described mouse fetal tracheal occlusion model (Aydin et al., 2018) to test whether the same increase in lung basal cells was seen in a different animal model of TO. After TO at E16.5, E18.5 fetal lungs were collected and single cell suspensions analyzed by flow cytometry. TO lungs had more than twice the number of basal cells at control lungs (Figure 2A-B). In a separate experiment, the lungs of TO mice had ∼2.5-fold greater quantity of the basal cell marker ketatin-14 (Figure 2C). Analysis of whole-lung sections of control and FETO mice revealed increased abundance and more distal extension of basal cells in TO lung compared to control (Figure 2D&E). These fetal mouse data collaborate findings in a fetal rabbit model that TO increases the number of basal cells.

**Figure 2:**
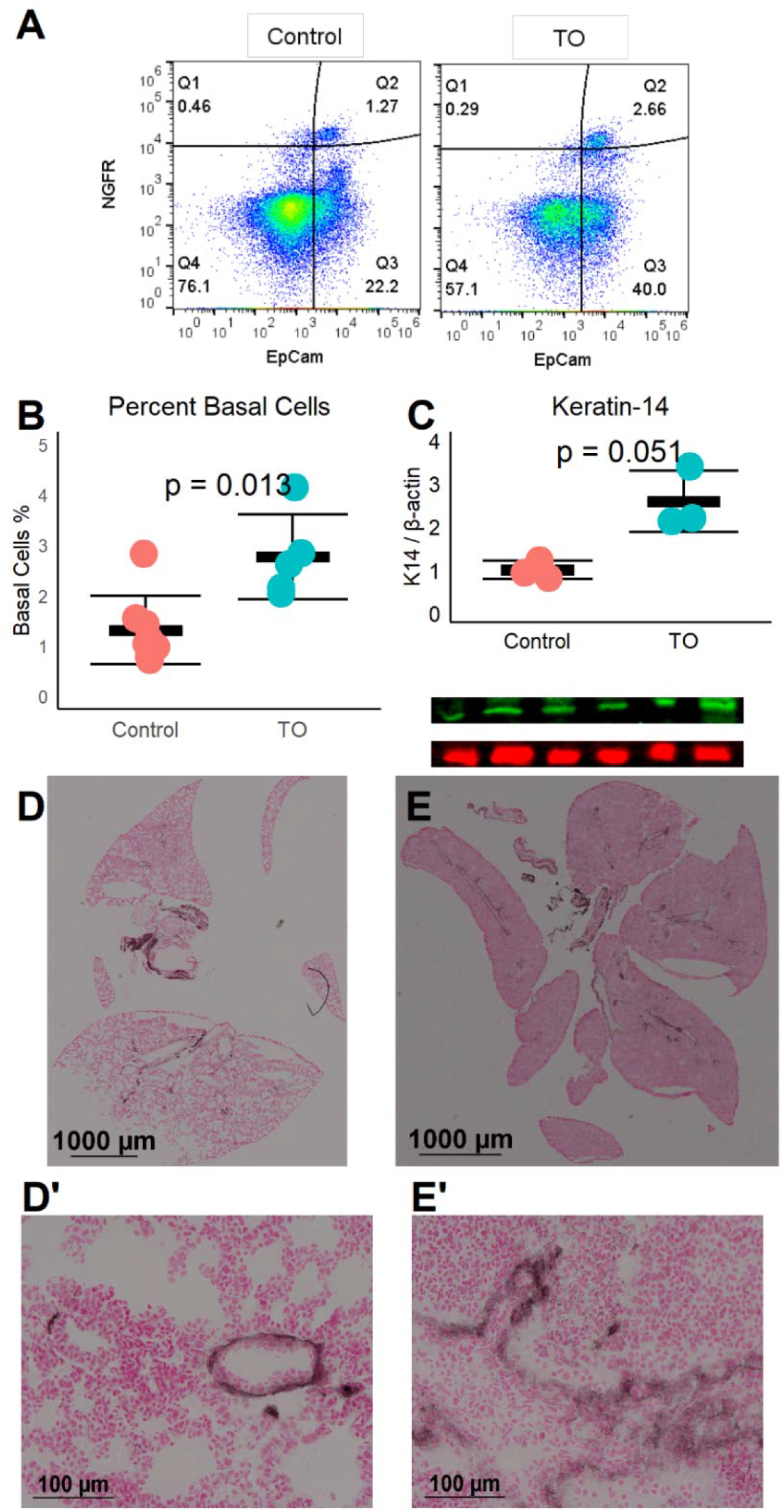
FETO Increases the Number and Distalization of Lung Basal Cells. (A) The percentage of lung basal cells as defined by co-expression of EpCam and Nerve growth factor receptor (NGFR) increased from ∼1.5% to ∼2.5% at E18.5 following TO at E16.5. (B) This increase percentage of basal cells in TO (n=6) compared to control (n=6) was consistent and statistically significant. (C) The whole lung protein content of the basal cell marker Keratin-14 was also increased. (D) Immunohistochemistry of control lung for NGFR showed restriction of basal cells to basement membrane of large conducting airways. (E) FETO lungs had increased cellularity and had extension of basal cells beyond the central conducting airways into the more distal conducting airways.

### Activation of Yap Signaling by FETO

The Yap pathway is both a regulator of epithelial cell proliferation and organ size and a direct regulator of p63, a necessary transcription factor for basal cells (Mahoney et al., 2014). Yap signaling is regulated by a series of kinases with phosphorylated Yap being targeted for degradation and non-phosphorylated Yap translocating to the nucleus where TEADs are recruited activating Yap-regulated genes (Meng et al., 2016). Thus, non-phosphorylated Yap is transcriptional active while phosphorylated Yap is sequestered in the cytoplasm. Immunohistochemistry for Yap and phospho-Yap identified grouped Yap-positive and phospho-Yap positive cells in E18.5 lung (Figure 3A-B). In contrast, in E18.5 FETO lungs, the majority of nuclei were Yap-positive and phospho-Yap staining was nearly absent (Figure 3C-D). Protein levels of the Yap-target connective tissue growth factor (CTGF) were increased in TO lung (Figure 3E). These data demonstrate the activation and transcriptional activity of Yap following TO.

**Figure 3:**
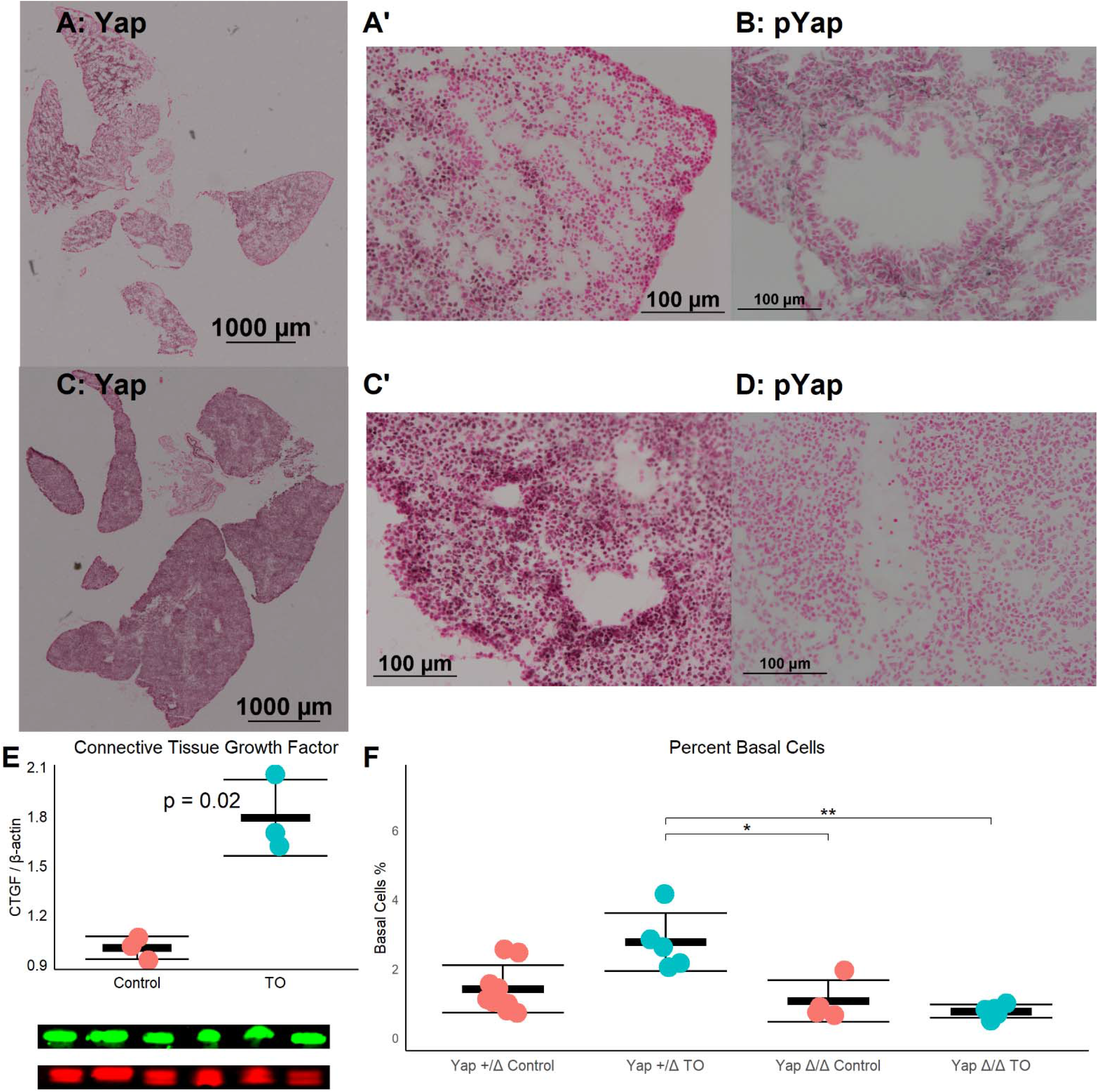
Increased Lung Epithelial Yap Signaling Is Responsible for Basal Cell Expansion. (A) In control E18.5 fetuses, non-phosphorylated (nuclear, active) Yap is present in small clusters of distal lung cells, and (B) phosphorylated (cytosolic, non-active) Yap is widely present. (C) In TO E18.5 lung, non-phosphorylated Yap is present more homogenously throughout the lung and can be detected in a more nuclei than in control with denser abundance of nuclear-Yap containing cells round airways. (D) In TO lung, phosphorylated Yap is less abundant than in control lung. (E) Western blot of lung homogenate showed increased levels of Connective tissue growth factor, a Yap-target gene. Comparison by Welch’s t-test. (F) Mice with hemizygous deletion of lung epithelial cell Yap at baseline (n=9), and after TO (n=5) showed similar numbers of basal cells as wild type mice. Lung epithelial cell-specific deletion of Yap without TO (n=4) or with TO (n=5) both had a numbers of basal cells comparable to control hemizygous and WT mice. *p<0.05, **p<0.01 by Holm-Sidiak post hoc test.

To test whether TO activation of Yap was directly responsible for basal cell expansion, we conditionally deleted Yap from the epithelium of fetuses using a Sonic hedgehog driver and floxed Yap alleles. There was a non-significant reduction in the number of basal cells between *Yap*^*LoxP/LoxP*^ or *Yap*^*+/*Δ^ mice and *Yap*^Δ^*/*^Δ^ at baseline and a significant reduction in the number of post-TO basal cells in *Yap*^Δ^*/*^Δ^ (Figure 3F). While these data cannot differentiate whether the lack of TO-induced basal cell expansion is from absence of a necessary progenitor or lack of a necessary transcriptional driver, they do show that Yap is necessary basal cell expansion in TO.

### Pathways and Processes Impacted by Tracheal Occlusion

We tested the relative weight of Yap transcriptional activity compared to other impacted signaling pathways by performing mRNA-seq and differential gene expression analysis in three control and three TO E18.5 lungs. TO lungs were most similar to each other (Figure 4A) although the separation between control and TO specimens was incomplete when assessed by the first two principal components (Figure 4B). After filtering for genes that contained at least 5 reads in three or more of the specimens, 17,374 genes were used in analysis. Among these genes, only 42 had adjusted p-values of <0.1. Gene set enrichment analysis (GSEA) of these genes identified inflammation-related, tube morphogenesis, and blood vessel morphogenesis processes as differentially regulated (Supplemental Table 2). For further analysis, we analyzed genes 2-fold increased (626 genes) or decreased (207 genes). Among upregulated genes, the biological processes that were activated were cardiac and striated muscle function-related, and there were no significant molecular functions identified among the downregulated genes (Supplemental Figure 2). Upstream analysis of genes with 2-fold change in expression identified interferons and toll-like receptors as activated, but it also identified regulators important in cell cycle and lung development such as TBX5, SPI1, CEBPA, TGFB1, AREG, FOXM1, and CCND1 (Figure 4C). Yap/Taz phosphorylation was predicted to be decreased (activation z-score -0.82, adjusted p-value = 1.49×10^−7^ and TEAD2 activity increased (activation z-score 1.63, adjusted p-value = 2.7 × 10^−7^) in this analysis, but the relative weights of these differences was smaller those of other regulators. Clustering of specimens using genes known to be regulated by Yap showed that TO and control specimens were distinct from one another (Figure 4D). In performing a similar analysis of lung cell-specific mRNAs as was done in fetal rabbit lung, basal cell, club/goblet cell, AT2 and myeloid cell mRNAs were increased in TO compared to control.

**Figure 4:**
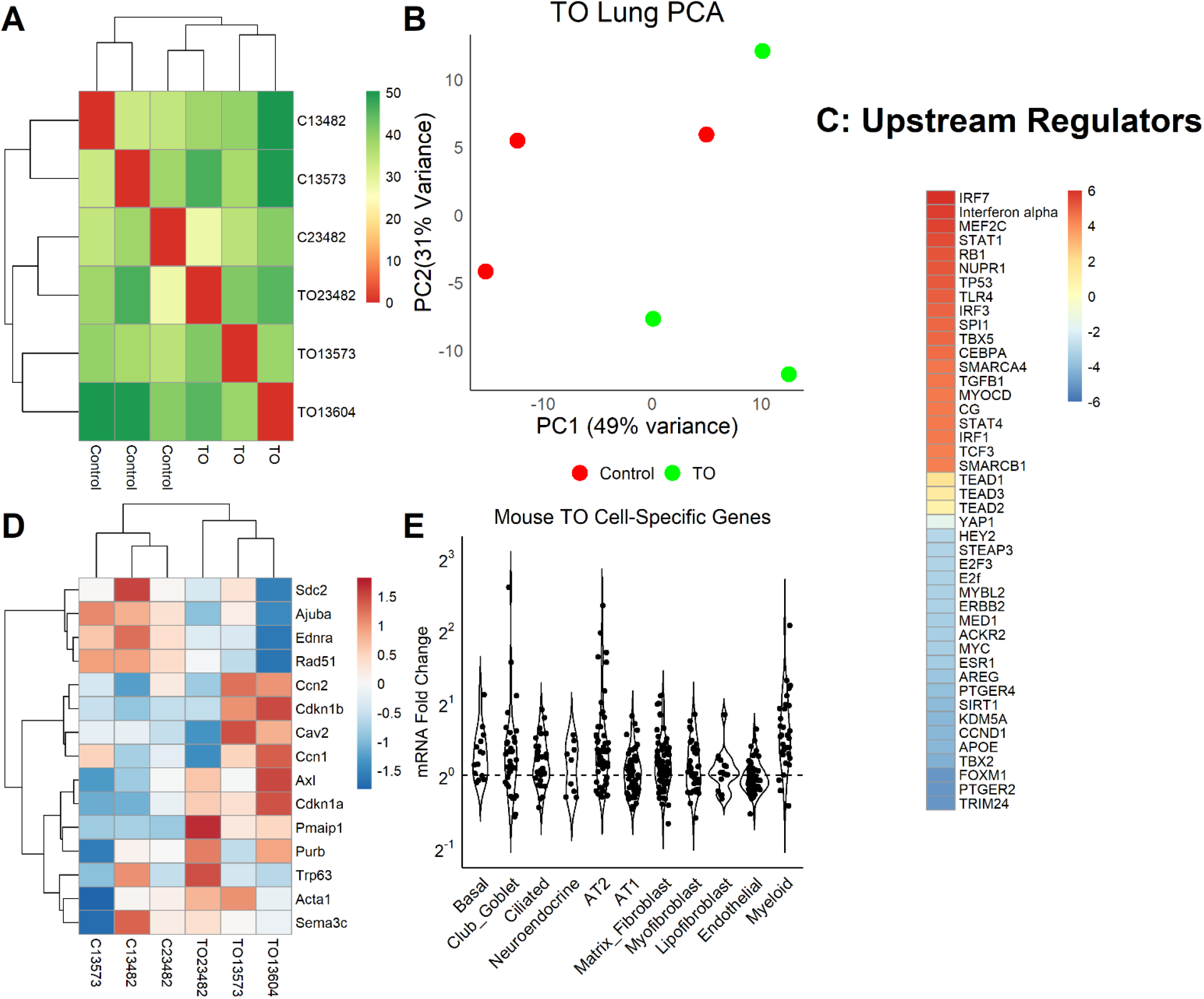
Yap in Mouse TO Lung (A) mRNA from the lungs of three control and three TO fetuses were analyzed by mRNA-seq. As assessed by sample distances, TO and control lungs were most similar to one another. (B) Principal component plot showed incomplete separation of control from TO specimens. (C) Ingenuity Pathway Analysis of upstream regulators of genes 2-fold changed between control and TO lungs showed that the regulators with the greatest predicted activation (red) were largely related to inflammation and the regulators with the greatest predicted inactivation (blue) were mostly related to cell cycle. Yap in transcriptionally inactive when phosphorylated, and non-phosphorylated YAP recruits TEAD factors to response elements. YAP and TEAD factors were not among the top 20 activated and inactivated regulators but were up and downregulated consistent increased nuclear Yap in TO. (D) Assessment of mRNA levels of genes known to be regulated by Yap were consistent with increased nuclear Yap in TO lung specimens. (E) Assessment of lung cell-specific mRNAs showed increased basal cell (1.17), club/goblet cell (1.15), AT2 (1.21), and myeloid cell (1.45) mRNAs in TO compared to control lungs.

### Human Tracheal Occlusion Results in Increased Yap and Extension of Basal Cells Distally

To determine whether Yap activation and distal lung basal cell expansion also occurred in human tracheal occlusion, we analyzed lung specimens from two infants that died shortly after birth with congenital high airway obstruction (CHAOS, gestational ages 34 and 36 weeks), three infants that died either before or shortly after delivery following endoscopic fetal tracheal occlusion (FETO) for congenital diaphragmatic hernia, and two lung specimens of infants who died of non-pulmonary causes at those same ages. All lung specimens had the typical position and spacing of basal cells in the major conducing airways (Figure 5A), and basal cells were not identified apart from these conducting airways in control lung specimens (Figure 5B). CHAOS lung specimens (Figure 5C&D) had clusters of basal cells in the airspaces of the distal lung with Yap-positive nuclei close to these cells. FETO lung specimens (Figure 5E&F) had expansion of basal cells in the distal airways; although, nuclear Yap staining was not prominent. These data are in agreement with finding in the mouse model of tracheal occlusion with regards to Yap activation and the presence of basal cells in the distal lung airspaces.

**Figure 5:**
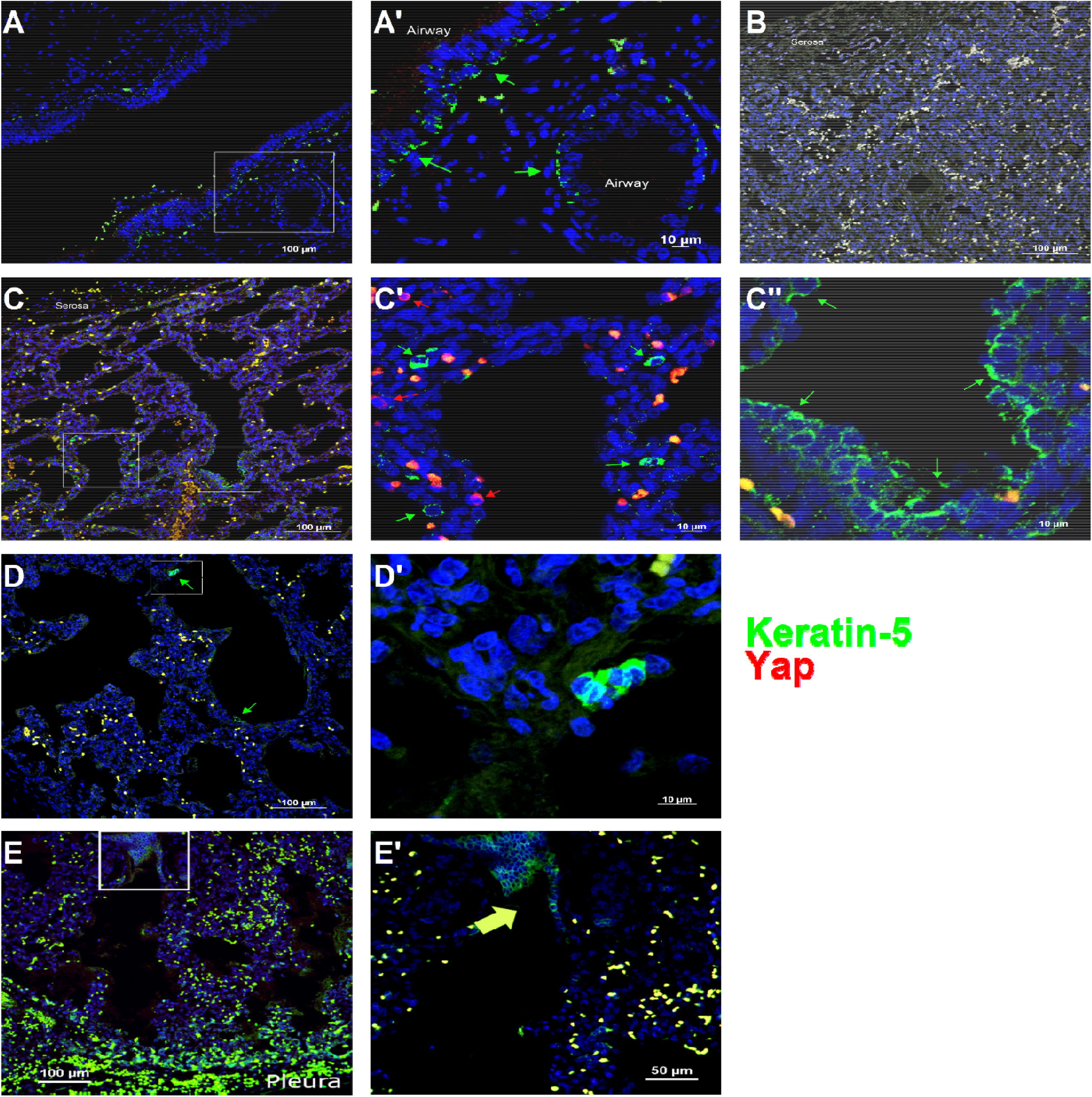
Basal Cells are Present in the Distal Lung of Infants with Congenital High Airway Obstruction. (A) As lung specimens from recently deceased infants that had undergone fetal tracheal occlusion were unavailable, we used lung specimens from infants gestational age 34-36 weeks that died within 48 hours of birth with and without congenital high airway obstruction (CHAOS). The lungs of both groups of infants had Keratin-5 positive cells (green arrows) beneath the pseudostratified epithelium of the large conducting airways. (B) The subpleural lung of control specimens had no Keratin-5 positive cells. (C) CHAOS lung had regions of lung with groups of Keratin-5 positive cells. None of these cells were Yap positive (red arrows) although cells containing both cytoplasmic and nuclear Yap were observed near Keratin-5 positive cells. (D) Another CHAOS lung specimen showing clusters of Keratin-5 positive cells just below the pleura. (E) Specimen of an infant with congenital diaphragmatic hernia treated with endoscopic fetal tracheal occlusion that died shortly after birth. In airways just below the pleura, clusters of keratin-5 positive basal cells can be identified.

## DISCUSSION

This is the first study to show changes in lung cell population distribution in response to FETO and is also the first to study the role of a specific signaling cascade in tracheal occlusion. While the role of Yap in specifying lung epithelial cell populations is well established, this study is the first to show its relevance in human lung development. To the best of our knowledge, this is also the first mechanistic study in fetal tracheal occlusion. To date, animal studies have been hampered by the need to use large animal models that are less amenable to genetic manipulation. The development of a minimally invasive mouse TO model (Aydin et al., 2018) allowed us to use a lung epithelial cell-specific deletion of Yap to mechanistically test its role in the observed expansion of basal cells in TO mouse lung.

Yap has a well-established roles in defining the proximal conducting airway epithelium in early lung development (Mahoney et al., 2014) and in maintaining lung basal cells in more mature lung (Yang et al., 2018). In the distal lung epithelium, cytoplasmic Yap plays an important role in AT2 cell and eventual AT1 cell differentiation. Yap that cannot undergo nuclear-cytoplasmic shuttling (i.e. sequestered in the nucleus) led to disorganization of the conducting airway epithelium without evidence of increased proliferation and with evidence of ectopic AT2 and AT1 cell markers (Soldt et al., 2019). However, Yap was also recently shown to mediate AT1 to AT2 cell conversion with an important role in post-injury repair (Penkala et al., 2021). Mst1 and Mst2 phosphorylate Yap leading to cytoplasmic sequestration. *Mst1* and *Mst2* deletion also impairs differentiation of conducting airway epithelial cells, but increases proliferation of these undifferentiated epithelial cells (Lange et al., 2015). In the Lange study, increased total and nuclear lab led to expansion of lung basal cells at the expense of club cells with this effect being mediated by the Yap target *Ajuba*. Interestingly *Ajuba* was decreased and *Trp63* levels mixed in TO lungs, perhaps reflecting counterregulatory mechanisms. We did not observe any increases in AT1 as was previously described in mice with deletion of *Lats1* and *Lats2* (which are downstream of *Mst1* and *Mst2* also leads to increased nuclear Yap). Interestingly, ectopic p63-positive cells were observed in *Mst1*&*Mst2*-deficient mice (Nantie et al., 2018). Our finding that a TO-mediated increase in nuclear Yap mediates expansion of the lung basal cell population is thus largely consistent with the literature on the role of Yap in lung epithelial cell differentiation and function with some differences likely attributable to differences in the four models (constitutive nuclear Yap, *Mst1*&*Mst2* deletion, *Lats1*&*Lats2* deletion, and TO with lung epithelial cell Yap deletion).

Clinically, little is known about the post-FETO lungs of infants with CDH. While FETO improves the survival of fetuses with the most severe forms of CDH (Al-Maary et al., 2016), impaired lung function is a substantial problem in survivors. The importance of basal cell clusters in the distal lung is also unclear. Basal cells or basal-cell progenitors have been shown to be contributors to distal lung regeneration following influenza infection (Yang et al., 2018), and Keratin-14-positive cell “pods” can be identified in the normal and regenerating adult distal lung (Vaughan et al., 2015). Increased percentages of basal cells are associated with several non-malignant pulmonary diseases when assessed by single-cell mRNAs-seq (Zuo et al., 2020). Our studies are not comprehensive enough to describe when Keratin-14 positive pods may appear in the human lung as we did not observe any in control lung specimens. Whether these cells persist or are adaptive or deleterious in the context of subsequent lung injury can now be assessed by removal of the tracheal ligature at E17.5 with subsequent parturition and raising of post-TO mice (Aydın et al., 2021b).

Although we focused Yap and basal cells, our bioinformatic data suggests that other signaling cascades are more significantly impacted than this one. The most notable difference between TO and control mouse lungs is increased inflammation in TO lung. The role of different lung resident macrophages in lung development is being increasingly appreciated (Tan and Krasnow, 2016), and how TO alters the relative abundance and activity of these populations is likely important in the post-TO lung. The TO lung also has activation of many cardiac and skeletal muscle-related pathways. This could be from three non-exclusive sources: lung myofibroblasts, vascular smooth muscle cells, and airway smooth muscle cells. Lung mesenchymal cells in general and lung myofibroblast in particular play an important role in saccular and alveolar lung development (Endale et al., 2017; Riccetti et al., 2020). Pulmonary vascular pruning, vascular smooth muscle hypertrophy, and pulmonary hypertension are substantial problems for infants with congenital diaphragmatic hernia (Coughlin et al., 2015). In rabbit models of CDH and TO, TO enhanced lung perfusion (Cruz-Martinez et al., 2009) and reduced arterial wall thickness after surgical DH (Prat Ortells et al., 2014). The potential impact of TO in airway smooth muscle hypertrophy and lung fibroblast populations merit further investigation which is now more achievable with mouse TO models. Third, TGF-β has been consistently reported as increased after TO in rabbit and ovine models (Peiro et al., 2018; Varisco et al., 2016; Vuckovic et al., 2016), and our study shows this to be true in the mouse model as well. While caution is required when making cross-species comparisons (Varisco et al., 2016), consistent findings across multiple should provide confidence that observed alterations are likely operative in humans.

Several study limitations are noted. For the quantification of lung cell populations in fetal rabbit lungs, quantitative image analysis of lung section is subject to measurement errors due to differences in tissue sections, sectioning plane, and imaging plane. The staining of all sections together, imaging, thresholding, and quantification using identical settings somewhat mitigate these concerns, and the correlation of image analysis and mRNA data provide confidence with regards to relative changes in cell populations between experimental group. However, the comparison of the relative abundance of one cell population to another should be made with caution. We quantified only the relative percentage of basal cells in TO lung. This is relevant because hypercellularity is known to occur following TO (Aydin et al., 2018; Varisco et al., 2016). We did not determine whether there were increased quantities of basal cell-associated mRNA and protein in TO basal cells, or if this increased expression was due solely to an increased number of basal cells. We did not explore the impact of TO on mesenchymal or immune cells although these signatures were more highly altered than that of Yap and basal cells. Finally, we could not obtain any lung tissues to directly assess basal cells in post-TO human lung, but we were able to secure a small number of specimens from infants with congenital occlusion of the trachea. Development of a biobank for CDH and FETO lung specimens of expired infants would greatly enhance the translatability of animal work being done in this area.

In summary, we have shown that Yap mediates the expansion of the basal cell population in mouse and human lung in response to tracheal occlusion.

## Data Availability

All data from this manuscript is freely available by contacting the corresponding authors.

## ACKNOWLEDGEMENTS

We would like to acknowledge the confocal imaging core and research flow cytometry core at Cincinnati Children’s Hospital Medical Center for technical assistance.

## SUPPLEMENTAL MATERIAL

### PCR Validation of Fetal Rabbit Lung mRNA-Seq Cell-Specific mRNAs

We quantified the mRNA levels of several cell-specific mRNAs in Control, CDH, TO, and CDHTO fetal rabbit lung. In general, the data showed good correlation of the cell-specific mRNAs with one another, a reduction in ciliated cell mRNAs, and increased basal cell and alveolar type 1 cell mRNAs (Supplemental Figure 1).

**Supplemental Figure 1:**
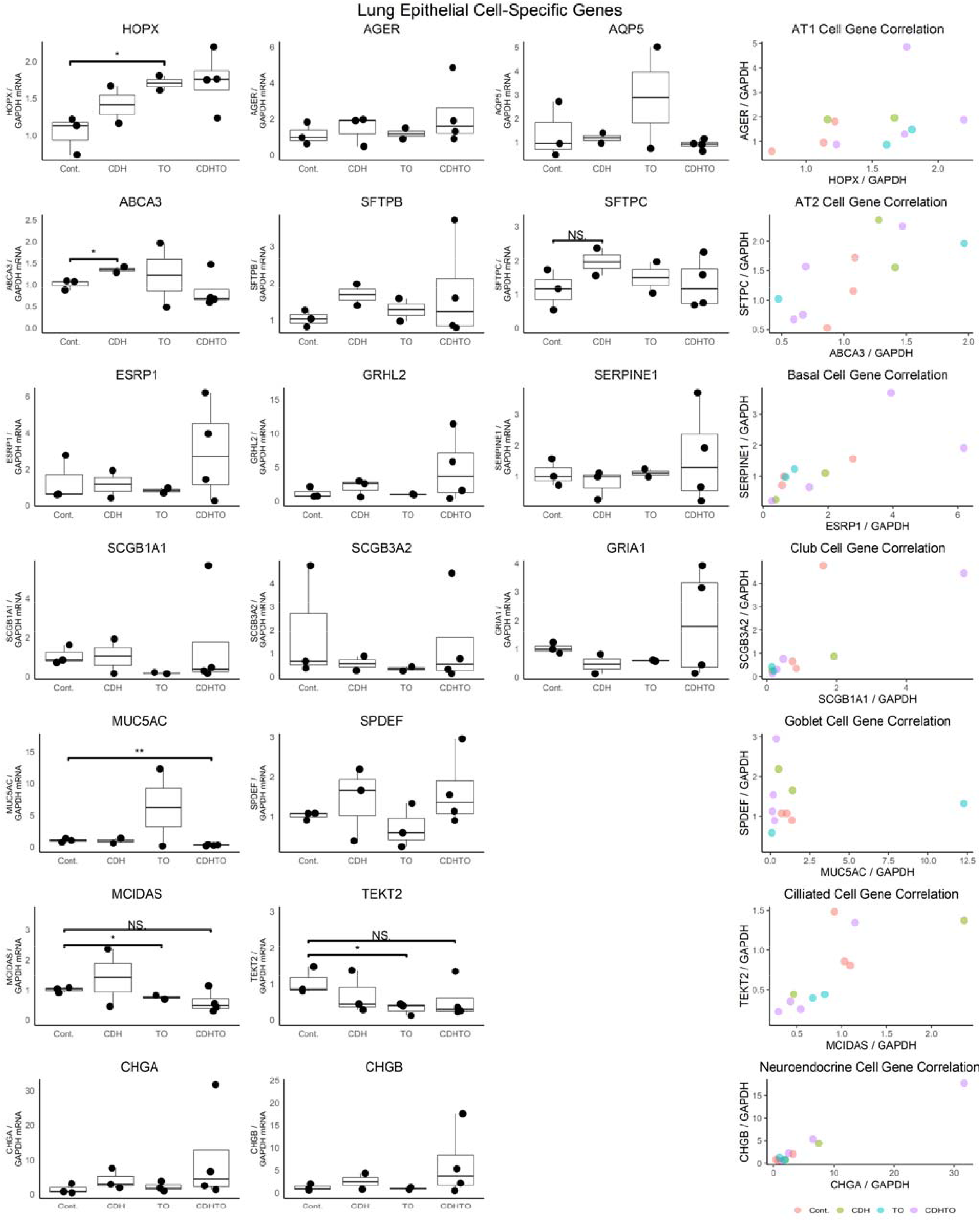
PCR quantification of cell-specific mRNAs in Fetal Rabbit lung. PCR for the AT1 cell markers Homeodomain-only protein homeobox (HOPX), Advanced Glycosylation End-products Receptor (AGER), and Aquaporin 5 (AQP5), the AT2 cell markers ATP Binding Cassette Subfamily A Member 3 (ABCA3), Surfactant Protein B (SFTPB), and SFTPC, the basal cell markers Epithelial Splicing Regulatory Protein 1 (ESRP1), Grainyhead Like Transcription Factor 2 (GHRL2), and Serpin Family E Member 1 (SERPINE1), the club cell markers Secretoglobin Family 1A Member 1 (SCGB1A1), SCGB3A2, and Glutamate Ionotropic Receptor AMPA Type Subunit 1 (GRIA1), the ciliated cell markers Multiciliate Differentiation And DNA Synthesis Associated Cell Cycle Protein (MCIDAS), and Tektin 2 (TEKT2), and the neuroendocrine cell markers Chromogranin A (CHGA) and CHGB were generally consistent with mRNA-seq data showing increased AT1 and basal cell mRNAs and decreased ciliated cell mRNAs.

### Cell-specific Genes

Genes in Supplemental Table 1 were used for deconvolution of bulk mRNA-seq data.

**Supplemental Table 1:**
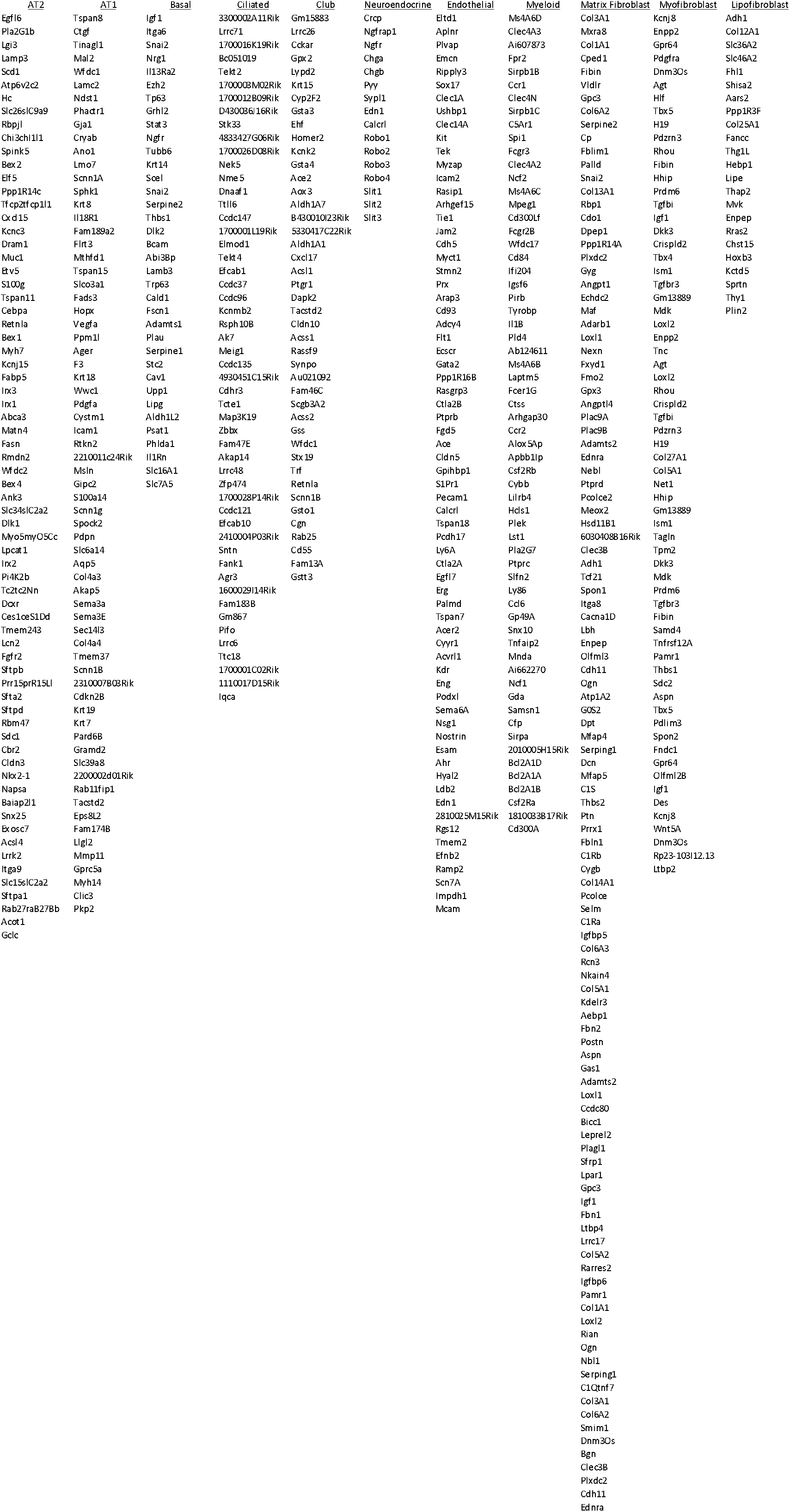
Cell-Specific Genes.

### Mouse TO mRNA-Seq Gene Set Enrichment Analysis

Significantly different, 2-fold upregulated, and 2-fold downregulated DEGs were analyzed in ToppGene to identify key pathways and processes as described in Supplemental Figure 2.

**Supplemental Figure 2:**
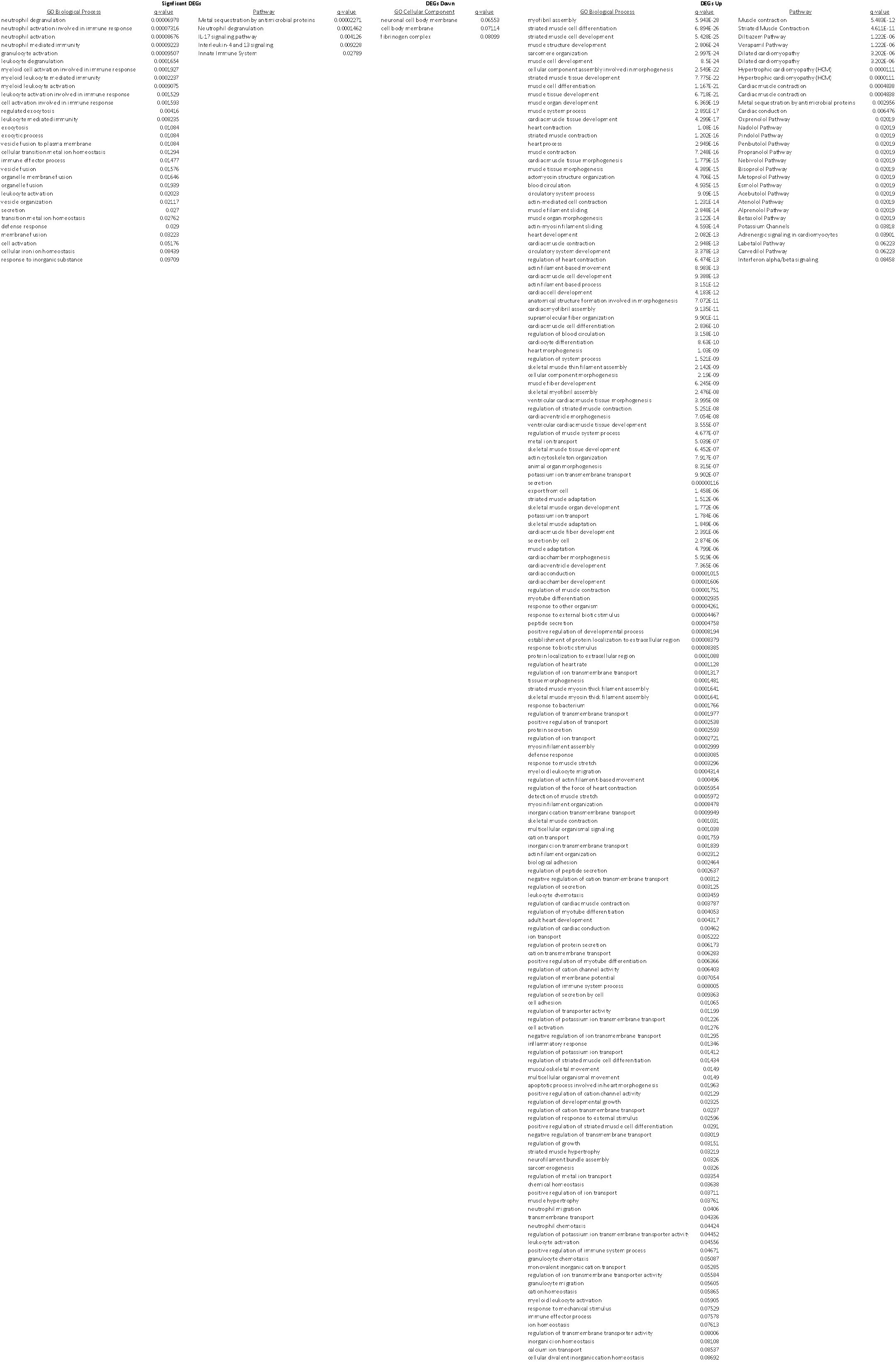
Gene Set Enrichment Analysis of DEGs in Mouse TO DEGs Down.

### Primers Used for Cell-Specific mRNAs in Rabbit Lung

The primers in Supplemental Table 3 were used with SybrGreen RT-PCR to quantify cell-specific mRNAs.

**Supplemental Table 3:**
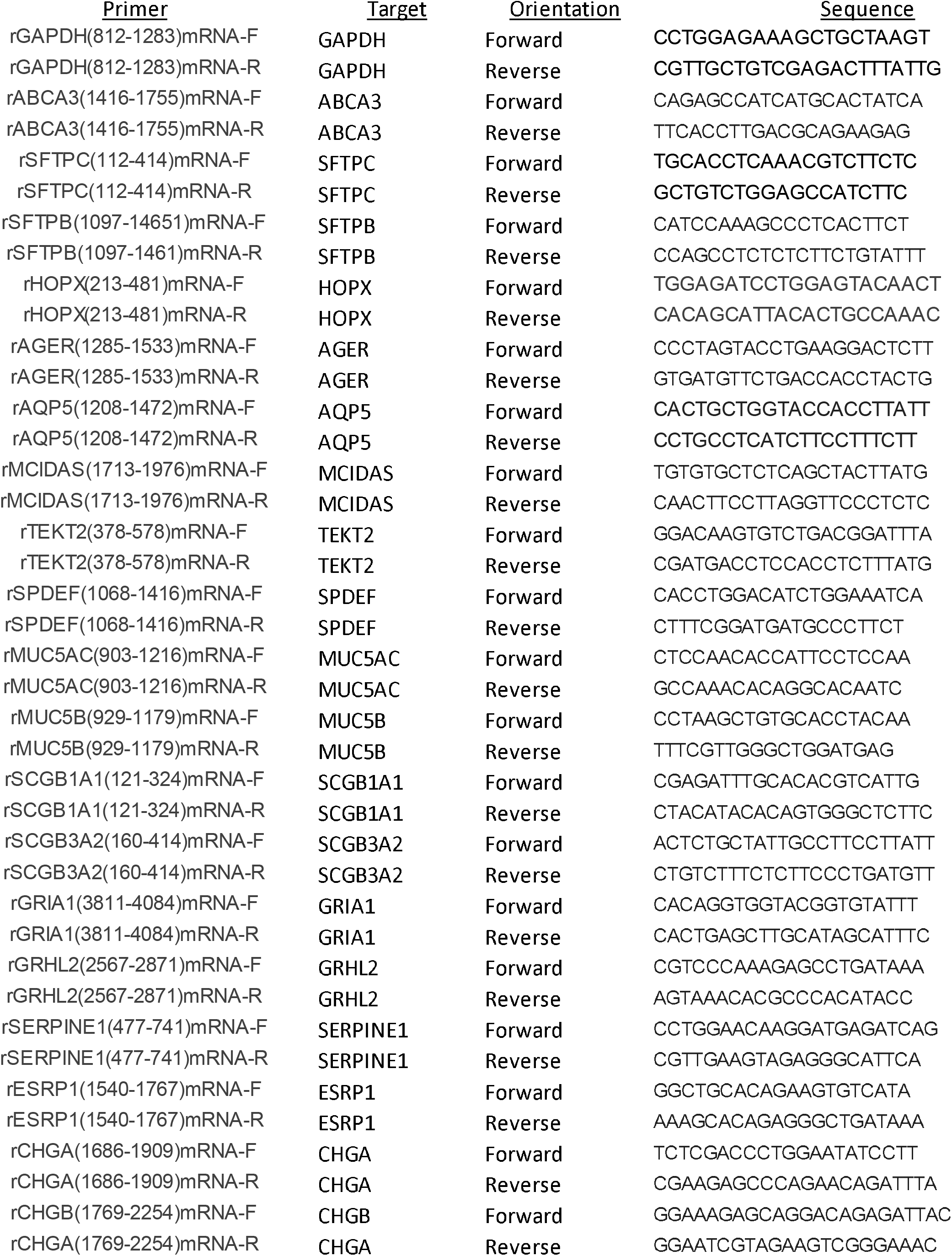
Primers for Quantification of Rabbit Lung-Cell Specific mRNAs

## Notes

### Competing Interest Statement

The authors have declared no competing interest.

### Funding Statement

Work was supported by NIH R01HL141229

### Author Declarations

Cincinnati Childrens Hospital Institutional Review Board (2016-9641) Cincinnati Childrens Hospital IACUC (2019-0034)

